# Efficacy and safety of Ayurveda interventions for Sinusitis: A systematic review and meta-analysis

**DOI:** 10.1101/2021.03.11.21253190

**Authors:** Azeem Ahmad, Manohar S. Gundeti, Parth P. Dave, Sophia Jameela, Shruti Khanduri, B.C.S. Rao, N. Srikanth

## Abstract

**Objectives:** To provide a broad evaluation of the efficacy and safety of Ayurveda interventions (procedural and non-procedural) for the management of sinusitis, and also of the relative efficacy and safety of different Ayurveda therapies for Sinusitis.

**Methods:** Five electronic databases for published research articles, three databases for the unpublished doctoral thesis, clinical trial registries, and hand searches were done till August 2020. All comparative clinical trials recruiting sinusitis patients of any age group, receiving Ayurveda intervention, regardless of forms, dosages, and ingredients, for not less than one week were included. The data extraction and the risk of bias(RoB) assessment were done by two reviewers independently.

**Results:** A total of 2824 records were identified, of which 09 randomized parallel arms trials met inclusion criteria. No studies were found comparing Ayurveda versus placebo or non-Ayurveda interventions. Combined Ayurveda therapy (CT) was statistically more beneficial compared with either procedural or non-procedural Ayurveda therapy alone in reducing symptoms nasal discharge (standardized MD −0.71, 95% CI −1.16 to −0.26, I^2^ 58%, 210 participants) and headache (standardized MD −0.44, 95% CI −0.86 to −0.02, I^2^ 56%, 218 participants), however, no significant difference was found in reducing symptoms nasal obstruction and loss of smell. No numerical data related to the safety of Ayurveda intervention was found in included trials. Because, included trials(09) were having ‘high’ to ‘unclear’ overall bias, sub-standard methodology, and heterogeneity in results, the overall findings need to be interpreted cautiously.

**Conclusions:** Although individual studies appeared to produce positive results, very low certainty of total effect(downgraded twice for RoB, once for inconsistency, indirectness, and imprecision each) hindered to arrive at any conclusion regarding efficacy or safety of Ayurveda interventions for sinusitis. There is a need for well-designed-executed-reported clinical studies on clinically relevant outcomes.

**PROSPERO registration number:** RD42018103995

**ARTICLE SUMMARY:** *Strength and limitations of this study:*

- This is the first systematic review to provide the status of available evidence on the efficacy and safety of Ayurveda interventions for sinusitis.
- The search strategy was comprehensive, all the relevant sources were searched for published as well as unpublished research works.
- This systematic review has a broad review question, which compromises its eternal validity.
- The certainty of the overall effect is ‘very low’ due to ‘unclear’ to ‘high’ overall risk of bias, lack of validated outcome measures, inconsistency in results with wide CIs, small sample sizes, incomplete reporting, etc.

## BACKGROUND

All the recently published guidelines have adopted the term rhinosinusitis (RS) instead of sinusitis. RS is the inflammation of the nose and paranasal sinuses characterized by two or more symptoms, one of which should be either, nasal blockage/obstruction/congestion or nasal discharge and the second either facial pain/pressure or reduction/loss of smell or both of these with objective findings on either computed tomography or nasal endoscopy[1]. In acute rhinosinusitis(ARC), there is complete resolution of symptoms within 12 weeks of onset, and persistence of symptoms beyond is categorized as chronic rhinosinusitis(CRS). ARC usually has an infective etiology while the etiology of chronic rhinosinusitis is likely to be multifactorial, with inflammation, infection, and obstruction of sinus ventilation playing a part[2].

CRS is an important chronic public health problem affecting the quality of life of more than 5% of people[3]. The overall prevalence of symptom-based CRS in the population has been found to be between 5.5% and 28%, while when symptoms are combined with endoscopy or CT scan prevalence is reduced to 3-6%[1]

The treatment strategy of RS includes oral and nasal antibiotics, steroids, antihistamines as well as nasal sprays, and saline irrigation in chronic as well as acute conditions[1,4]. More than 1 in 5 antibiotics prescribed in adults are for RS, making it the fifth commonest diagnosis liable for antibiotic therapy[4]. Emerging threats of antibiotic resistance have necessitated the need for exploring other interventions that could offer better or comparable efficacy and safety.

Ayurveda is one among various Indian Traditional systems of medicine which have been practiced in India for centuries. The clinical features of the disease *Pratishyaya* mentioned in different Ayurveda texts have symptomatic similarity to sinusitis/rhinosinusitis[5]. Depending upon the characteristics of the disease, and pathophysiological characteristics of the patient, treatment strategies are designed in Ayurveda. Modalities of treatment include both *Shodhana* (bio-purificatory therapies including *Panchakarma*) and *Shamana* (palliative therapy). Ayurveda standard treatment guideline was published by the Ministry of AYUSH, Government of India, which recommends different Ayurveda treatment strategies and modalities according to the different clinical settings[6]. However, the lack of empirical evidence and biological plausibility of the probable efficacy or safety of the interventions enlisted for clinical use probes the gathering of evidence through systematic review and meta-analysis. It was concluded in a systematic review that “There is no evidence of benefit from antibiotics for the common cold or for the persisting acute purulent rhinitis in children or adults. There is evidence that antibiotics cause significant adverse effects in adults when given for the common cold and in all ages when given for acute purulent rhinitis”[7]. If the review could gather conclusive evidence, on the safety and efficacy of Ayurveda interventions, that could effectively manage the disease without resorting to antibiotics, it would be highly beneficial to the patients. Since there is a paucity of robust, clinically oriented scientific studies in Ayurveda[8,9], this would at least aid in identifying the lacunae in available evidence and would pave the way for conducting robust clinical studies. Therefore, this systematic review was conducted to provide a broad evaluation of the efficacy and safety of Ayurveda interventions (procedural and non-procedural) for the management of sinusitis/rhinosinusitis and also to review the relative efficacy and safety of procedural therapy(*Shodhana)*, non-procedural therapy(*Shamana)*, and a combination of these for the management of sinusitis.

## METHODS

This systematic review was conducted following the guidelines of the Cochrane Handbook of Systematic Review of Interventions[10], and is reported as per the Preferred Reporting Guideline for Systematic Review and Meta-Analysis (PRISMA) guidelines[11], and the completed checklist is available as file S1. The protocol was registered prospectively with PROSPERO and available online as file S2.

### Eligibility Criteria

Studies fulfilling the criteria of **1. Study design:** All comparative clinical trials including randomized controlled trials (RCTs), quasi-randomized controlled trials, Non-randomized trials (nRCTs), multiple arms clinical trials. **2. Population:** Patients diagnosed with either sinusitis/rhinosinusitis (either diagnosed clinically or also confirmed with laboratory and radiological findings), or Pratishyaya/Pinasa as defined in Ayurveda, irrespective of age and sex. **3. Intervention:** Ayurveda treatment (*Shamana* or/and *Shodhana*) as standalone or add-on therapy with any dose, type, schedule, medicine, dosage form, either alone or combination, with or without *Pathya-apathya* (diet and lifestyle regimen). Patients may receive additional non-Ayurveda intervention in all groups of study. **4. Comparator:** The comparator arm utilizing Ayurveda interventions with a different dose, type, schedule, dosage form compared to intervention; or Non-Ayurveda interventions including contemporary interventions, Placebo, Sham therapy; or their combinations. **5. Duration of intervention:** Not less than one week.

### Outcomes of interest

The primary outcomes of interest are the response in terms of improvement in Subjective and/or objective criteria of assessment in RS and reported serious adverse events(SAE) resulting in death, disability or incapacity, life-threatening complications, that required hospitalization. Secondary outcomes included withdrawals of the participant from the study due to adverse events(AE)/adverse drug reaction(ADR), non-response to treatment or inconvenience of therapy/treatment, and the number of patients with specific AE.

### Study identification

We searched various databases including PubMed(Central), Cochrane Central Register of Controlled Trials(CENTRAL), AYUSH Research Portal (Govt of India), DHARA, Google Scholar, and Online clinical trial registers (CTRI, Clinicaltrials.gov & WHO-ICTRP). Furthermore, reference lists of related publications were also searched to get relevant publications. For unpublished postgraduate (P.G.) and doctoral (Ph.D.) dissertation works, we have searched the *Shodhganga* portal, university/Institutional website, and other potential sources from conception to August 2020, and studies reported in English or Hindi language were selected. The main search items included, ‘Ayurved*’, ‘sinusitis’, ‘Rhinosinusitis’, ‘Pratishyaya’, ‘Pinasa’, and their synonyms. Search strategy for AYUSH Research Portal and Cochrane CENTRAL databases are available online as file S3.

### Selection of studies and Data extraction

Two reviewers independently screened the title and abstract of identified articles. Potentially eligible articles were thoroughly scanned fully to match with eligibility criteria. If needed, the corresponding authors were contacted for additional information through e-mail or telephone.

Data were extracted for data items viz. authors name, year of publication, diagnosis, sample size, interventions, controls, safety and efficacy outcomes measures, follow-up, AE/ADR, and dropout with reason. Any disagreement was consulted and settled through discussions with a third reviewer, where necessary.

### Assessment of risk of bias and overall quality of evidence

Two reviewers independently assessed the risk of bias(RoB) according to the Cochrane RoB2 tool[10,12]. The assessment was made on the study level and any disagreement was resolved through discussion.

To assess the overall quality of evidence for primary outcomes, the Grading of Recommendations Assessment, Development, and Evaluation(GRADE) approach was applied with the help of an online available tool[13,14].

### Data Analysis

For continuous outcomes, the mean change from baseline for each group with corresponding standard deviations (SDs) was recorded, and for dichotomous data, the Risk ratio (RR) was calculated with a confidence interval of 95%. We used Review Manager Software (RevManV.5.3) for analysis[15], and got a pooled estimate of treatment effect as standardized mean difference(SMD) between groups and corresponding 95% CIs because in some included studies there was no information about scales used for measurement. We found diversity in interventions, controls, time point of assessment, and population, a random effect model was applied in all meta-analyses. The *I*^*2*^ statistic was used to calculate the statistical heterogeneity, and the value, greater than 60% was considered as significant, and sources of heterogeneity were analyzed. We performed a subgroup analysis where two categories of Ayurveda interventions in the control group i.e. Non-procedural therapy (*Shamana therapy)* or Procedural therapy (*Shodhana therapy)* were used. For sensitivity analysis, the “leave-one-out” method was used to assess the effect of individual study.

### Risk of bias across studies

All efforts were made to retrieve and compare the original trial protocols with the final publications and wherever possible, to identify any outcomes that were measured but not reported.

## RESULTS

### Study Selection

A total of 2824 records were identified from all sources. After the removal of duplicates and other irrelevant results, 68 records were available. After screening titles and abstracts, 44 records were excluded and the remaining 24 were further subjected to full-text screening as per the selection criteria. After the screening, 9 were included in qualitative analysis, among which 6 were subjected to meta-analysis (Fig 1).

**Fig 1.**
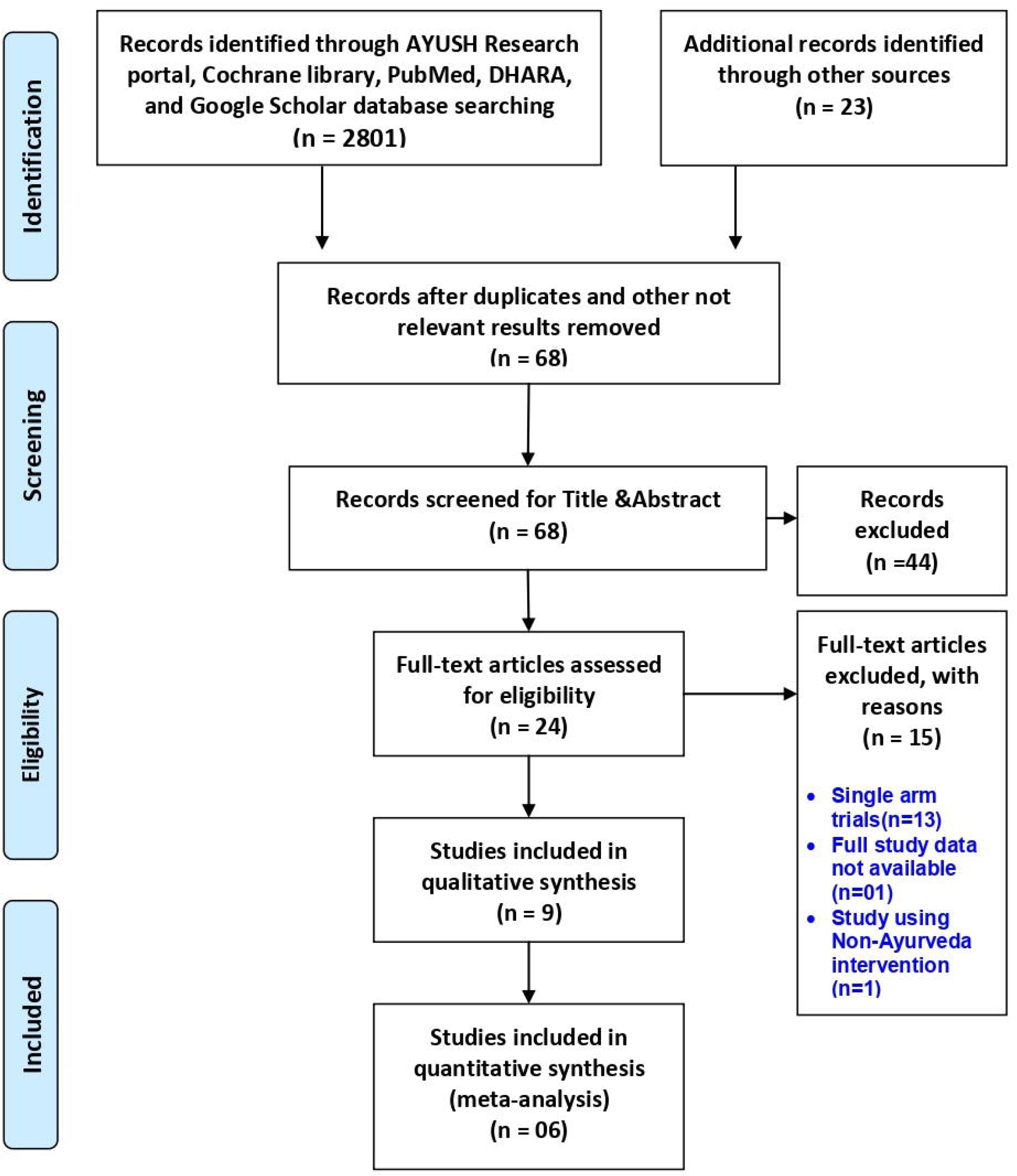
PRISMA flow diagram showing steps followed during study selection process. From: Moher D, Liberati A, Tetzlaff J, Altman DG, The PRISMA Group (2009) Preferred Reporting Items for Systematic Reviews and Meta-Analyses: The PRISMA Statement. *PLOS Medicine* 6(7): e1000097. doi:10.1371/ journal.pmed.1000097.

### Description of studies

Among these 9 randomized parallel arms trials (pre-treatment n = 307; end of treatment n = 279) [16-24], 6 trials were included in quantitative analysis (pre-treatment n = 287; end of treatment n = 259). Among them, 4 trials have multiple arms[16,18,24]. The mean participants in each arm of the trials were found to be 14. All the selected studies were conducted in India. All the trials compared different Ayurveda interventions parallelly and reported either partial or complete resolution of symptoms at specific time points. Subjective outcomes used were in the form of self-developed assessment scales according to the severity of the symptoms, ranging from 0 to 4, where 4 means worse state and 0 means no symptoms. In 4 trials, narrative reporting on adverse events was available[16,17,20,21]. Key data points from the included trials are presented in Fig. 2.

**Fig 2:**
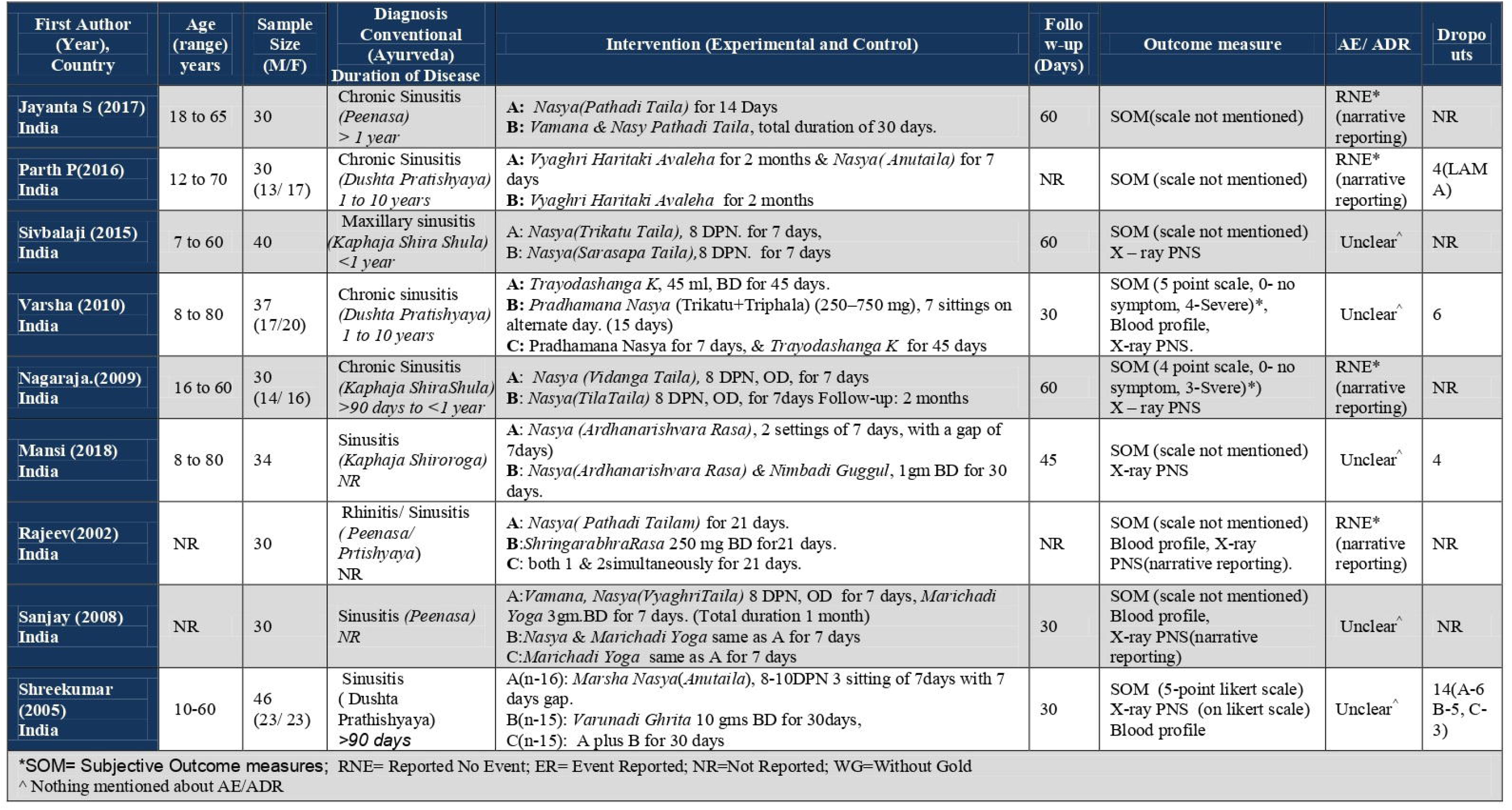
Characteristics of included trials in this systematic review and meta-analysis.

### Interventions

Three major categories of Ayurveda intervention were identified during the screening of search results which include 1. A combination of Ayurveda procedural & non-procedural interventions (CT), 2. Ayurveda procedural interventions (PT) alone, and 3. Ayurveda non-procedural oral interventions (NPT) alone. PT and NPT collectively reported as Single Therapy(ST) in this review. Details regarding the interventions used in the included studies are available online as file S4.

### Risk of bias within studies

Generally, the quality of the included studies was poor due to unclear or high overall risk of bias(Fig.3). In most of the included trials, it is only mentioned that “participants were randomly allocated into the groups” and no further details about the randomization process were provided, and also there was no information about allocation sequence concealment and blinding process. Only one trial has reported the use of a random number table for random sequence generation[22], and the blinding was not reported in any trial. The reporting of outcome data (baseline characteristics, age of the participants, the number of participants included in each analysis, the number of participants who completed the treatment, and dropped outs) was also incomplete. Selective outcome reporting was not evaluated because we could not find registered protocols of all the included studies. In addition, the sample size of all the trials was small and none of the trials reported how sample size was calculated. It was also not reported in any of the trials that intention-to-treatment or per-protocol analysis was used and it was suspected that dropouts were excluded from the analysis.

**Fig 3.**
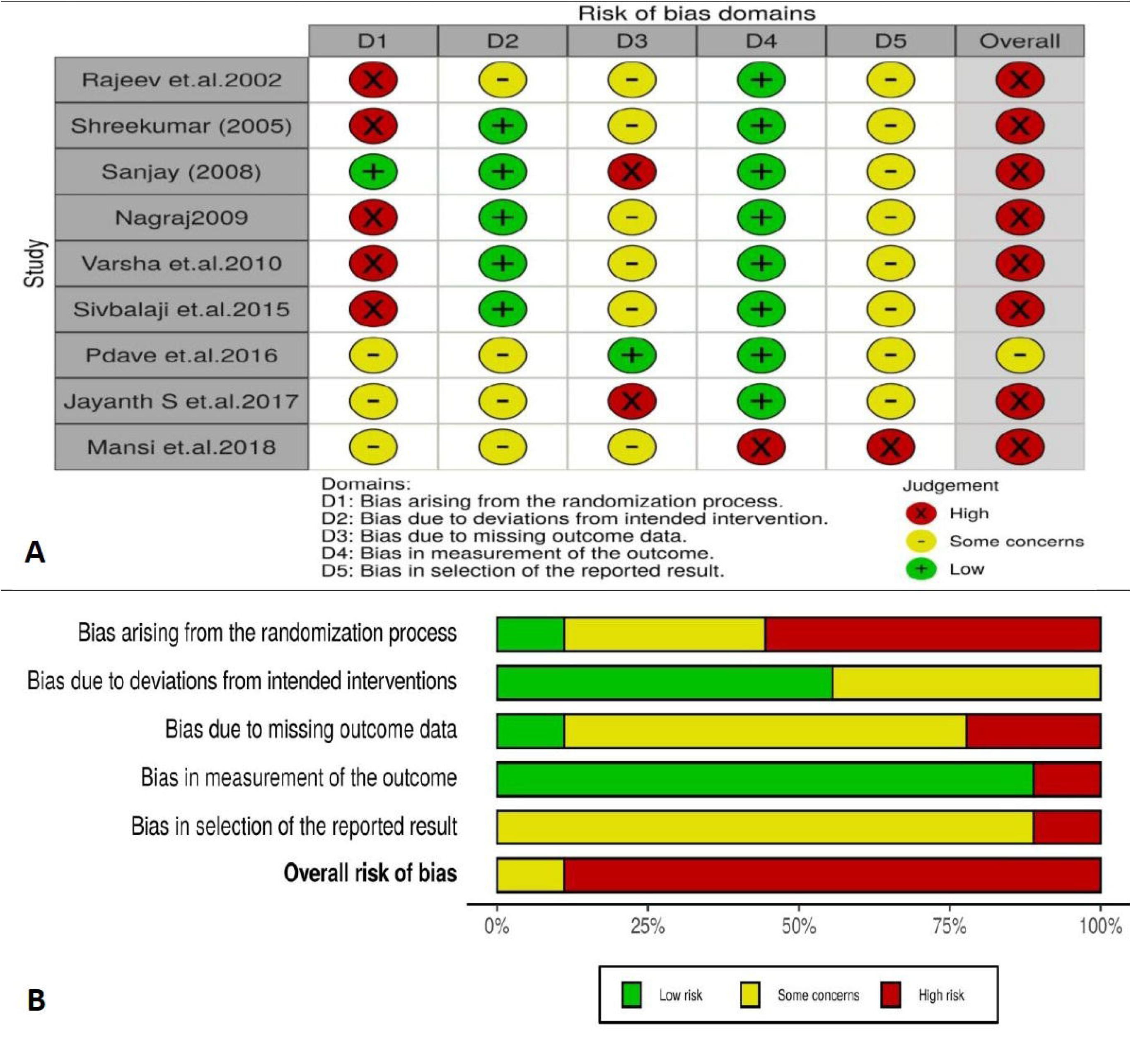
‘Risk of bias’ summary of randomized trials.

### Outcomes and Effectiveness of Ayurveda Interventions

As there were multiple subjective outcome measures of efficacy (signs and symptoms) were used within the study and which were also heterogeneous among the studies. Hence, we have selected the most common signs and symptoms reported in the selected studies viz. Nasal blockage/obstruction/congestion, nasal discharge, and reduction/loss of smell for meta-analysis analysis, which were included in the diagnostic criteria of RS suggested in the European position paper on rhinosinusitis and nasal polyps-EPOS 2012[1], and also the symptom “headache” which was assessed in most of the included studies.

### Comparison 1. CT versus ST

Data from 06 trials were pooled to determine the effects of CT versus ST strategies on the changes in symptom nasal discharge[15,17,19,21-23]. For 3-arm trials[16,18,24], we analysed each comparison separately(including 10 comparisons, total n=210).

Compared with ST, CT has a statistically significant positive difference in reduction in symptom nasal discharge (*Nasasrava*) (SMD = −0.71; 95% CI = −1.16, −0.26; p=0.002, *I*^*2*^=58%) after combining the data from 06 trials (including 10 comparisons, total n=210)[16,18,20,22-24], also in subgroup analysis (CT versus NPT) CT was found beneficial (SMD= −0.89; 95% CI = −1.53, −0.25; p=0.007, *I*^*2*^=69%), while the difference between CT and PT was statistically non-significant (SMD= −0.38; 95% CI = −0.88, 0.13; p=0.15, *I*^*2*^=0%) (Fig 4A).

**Fig 4.**
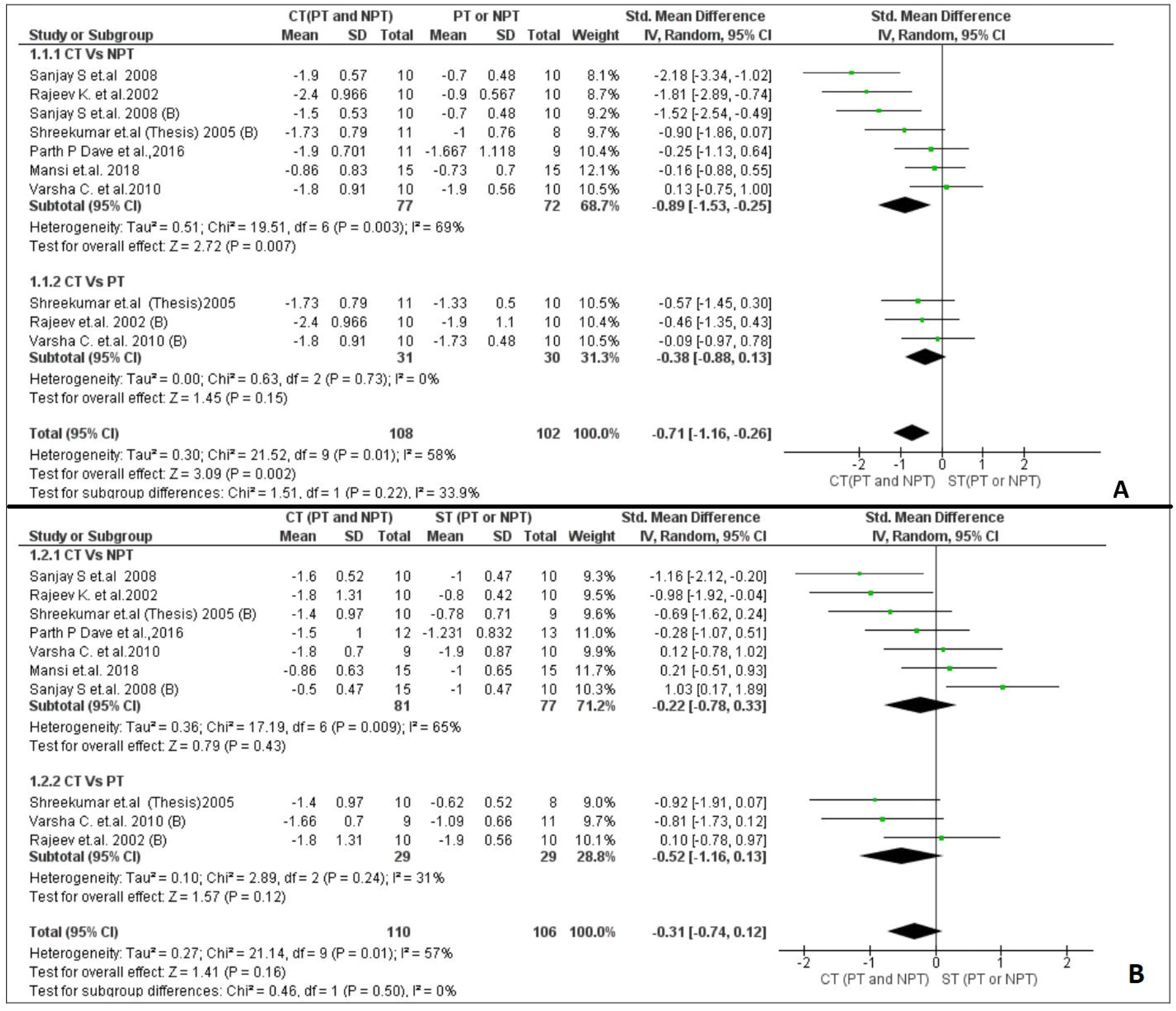
Forest plot of comparison: 1 Combined Therapy (CT) Vs Single Therapy (ST), outcome: 1.1 Nasal discharge (*Nasasrava*). Fig 4B. Forest plot of comparison: 1 Combined Therapy (CT) Vs Single Therapy (ST), outcome: 1.2 Nasal obstructions (*Nasa-Avrodha*).

Non-significant difference was found between CT and ST (SMD= −0.31; 95% CI = −0.74, 0.12; p=0.16, *I*^*2*^=57%) in reducing symptom nasal obstruction (*Nasa-Avrodha*), after combining the data from 06 trials (including 10 comparisons, total n=216)[16,18,20,22-24], also in subgroup analysis, the differences between CT and PT (SMD= −0.52; 95% CI = −1.16, 0.13; p=0.03, *I*^*2*^=31%), and CT and NPT (SMD= −0.22; 95% CI = −0.78, 0.33; p=0.43, *I*^*2*^=65%), were non-significant (Fig 4B).

CT was having a significant positive difference (SMD= −0.44; 95% CI = −0.88, −0.02; p=0.04, *I*^*2*^=56%) on reducing symptom headache (*Shirashoola*) after combining the data from 06 trials (including 10 comparisons, total n =218) [16,18,20,22-24], however in subgroup analysis, CT versus NPT, CT was found beneficial (SMD= −0.55; 95% CI = −1.06, −0.05; p=0.03, *I*^*2*^=57%), while analysing CT versus PT, the difference was statistically non-significant (SMD= −0.16; 95% CI = −1.02, 0.71; p=0.72, *I*^*2*^=64%) (Fig 5A).

**Fig 5A.**
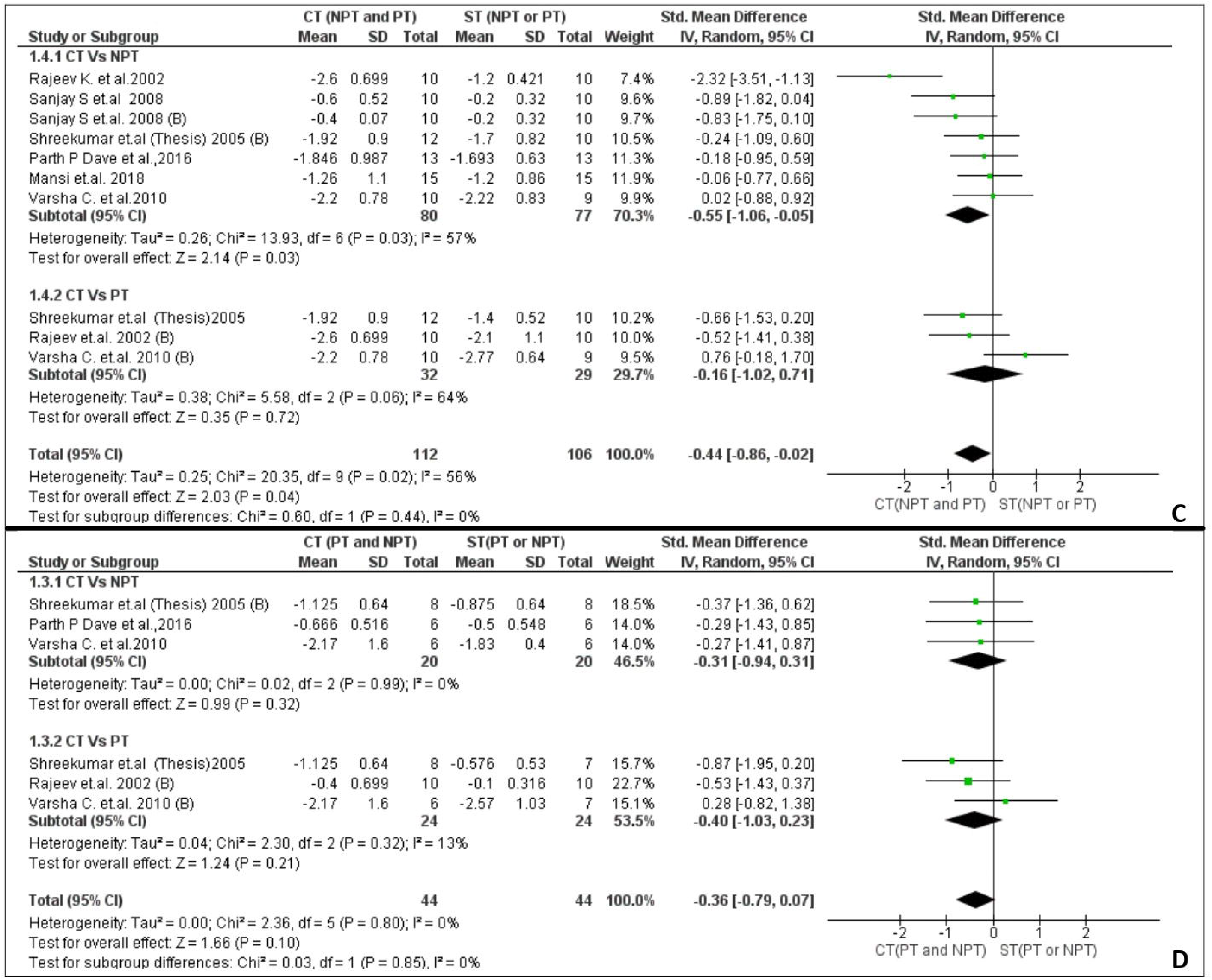
Forest plot of comparison: 1 Combined Therapy (CT) Vs Single Therapy (ST), outcome: 1.3 Headache (*Shirashoola*). Fig 5B. Forest plot of comparison: 1 Combined Therapy (CT) Vs Single Therapy (ST), outcome: 1.4 Loss of smell (*Ghrana-viplava*).

CT was having an insignificant difference in results (SMD= −0.36; 95% CI = −0.79, 0.07; p=0.10, *I*^*2*^=0%) on reducing symptom loss of smell (*Ghrana viplava*) after combining the data from 04 trials (including 6 comparisons, total n =88) [16,18,20,24], also no significant differences were found in both subgroup analysis, CT versus NPT (SMD= −0.31; 95% CI = −0.94, 0.31; p=0.32, *I*^*2*^=0%), and CT versus PT (SMD= −0.40; 95% CI = −1.03, 0.23; p=0.21, *I*^*2*^=13%) (Fig 5B).

### Comparison 2. PT versus NPT

Data from 03 trials [16,18,24] were pooled to determine the effects of PT versus NPT on symptoms of nasal discharge, nasal obstruction, loss of sense of smell/taste, and headache. There was no significant difference found between these two Ayurveda treatment strategies. Details are provided in the online file S5.

### Adverse events

None of the trials included any safety outcome measures in the assessment criteria. Also, none of the trials reported risk of AE /ADR in procedural and non-procedural Ayurveda interventions numerically; however, 4 trials reported ‘no events’ narratively in very short[17,18,21,24]. Only 4 studies reported dropouts[19,21,23,25], but only one of these reported a reason for this as LAMA (left against medical advice)[20]. Because the reasons for this attrition were not reported, it is difficult to exclude the chance of non-compliance due to unpalatability of Ayurveda interventions, discomforts with different therapeutic procedures, and unrecorded AEs/ADRs/SAEs.

## Discussion

### Summary of main results

We did not find any trials comparing Ayurveda interventions with non-Ayurveda interventions or placebo. However, we found trials testing the effects of two major modalities of Ayurveda, PT, and NPT in this disease condition. As per the protocol, we compared the combined effects of these two therapeutic modalities of Ayurveda against either of these, we found that the CT was statistically more beneficial in reducing the symptom headache (*Shirashoola*) and nasal discharge (*Nasa-shrava*). There was no significant difference found in reducing symptom nasal obstruction (*Nasa-avrodha*) and loss of smell (*Ghrana viplava*). We also compared the effect of PT with NPT, and no significant difference was found in the improvement of any of the above-mentioned symptoms. None of the trials have reported any data on few clinically relevant outcomes viz. safety outcome measures, AE, ADR, SAE, disease-specific quality of life, overall improvement, and economic outcomes. We are unable to make any conclusion or recommendation about these outcomes in all categories of comparisons. Details are given in Table 1, comparison 1 & 2.

**Table 1.** Summary of Findings (SoF) Table.

| Outcomes | № of participants (studies) | The certainty of the evidence (GRADE) | Relative effect (95% CI) | Anticipated absolute effects (95% CI) |  |
| --- | --- | --- | --- | --- | --- |
|  |  |  |  | Risk with Control | Risk difference with Intervention |
| Comparison 1: Aurveda CT versus ST |  |  |  |  |  |
| Nasal discharge (*Nasa-shrava*)<br>follow up: range 07 days to 60 days <sup>a,b</sup> | 210 (10 RCTs) <sup>c</sup> | ⊕□□□<br>VERY LOW <sup>d,e,f,g,h</sup> | - | The mean Nasal discharge ranged from - **1.9 to -0.7** | SMD **0.71 SD lower**<br>(1.16 lower to 0.26 lower) |
| Nasal obstruction (*Nasa-avrodha*)<br>follow up: range 07 days to 60 days <sup>a,b</sup> | 216 (10 RCTs) <sup>c</sup> | ⊕□□□<br>VERY LOW <sup>d,e,f,g,h</sup> | - | The mean Nasal discharge ranged from - **1.9 to -0.62** | SMD **0.31 SD lower**<br>(0.74 lower to 0.12 higher) |
| Loss of smell (*Ghrana viplava*)<br>follow up: range 07 days to 60 days <sup>a,b</sup> | 88 (6 RCTs) <sup>c</sup> | ⊕□□□<br>VERY LOW <sup>d,e,f,g,h</sup> | - | The mean Nasal discharge ranged from - **2.57 to -0.1** | SMD **0.36 SD lower**<br>(0.79 lower to 0.07 higher) |
| Headache (*Shirashoola*)<br>follow up: range 07 days to 60 days <sup>a,b</sup> | 218 (10 RCTs) <sup>c</sup> | ⊕□□□<br>VERY LOW <sup>d,e,f,g,h</sup> | - | The mean Nasal discharge ranged from - **2.77 to -0.2** | SMD **0.44 SD lower**<br>(0.86 lower to 0.02 lower) |
| Adverse Events (related to procedural Therapies) | No trial reported on outcome “Adverse Events (related to procedural Therapies)” |  |  |  |  |
| Other Adverse events | No trial reported on the outcome “Other Adverse events” |  |  |  |  |
| Comparison 2: Ayurveda PT versus NPT |  |  |  |  |  |
| Nasal discharge (*Nasa-shrava*)<br>follow up: range 07 days to 60 days <sup>a,b</sup> | 58 (3 RCTs) | ⊕□□□<br>VERY LOW <sup>d,e,f,g,h</sup> | - | The mean Nasal discharge ranged from - **1.9 to -0.9** | SMD **0.41 lower**<br>(1.22 lower to 0.4 higher) |
| Nasal obstruction (*Nasa-avrodha*)<br>follow up: range 07 days to 60 days <sup>a,b</sup> | 58 (3 RCTs) | ⊕□□□<br>VERY LOW <sup>d,f,g,h,i</sup> | - | The mean Nasal discharge ranged from - **1.9 to -0.78** | SMD **0.26 SD lower**<br>(2.01 lower to 1.48 higher) |
| Loss of smell (*Ghrana viplava*)<br>follow up: range 07 days to 60 days <sup>a,b</sup> | 28 (2 RCTs) | ⊕□□□<br>VERY LOW <sup>d,f,g,h,i</sup> | - | The mean Nasal discharge ranged from - **1.83 to -0.87** | SMD **0.16 lower**<br>(1.46 lower to 1.14 higher) |
| Headache (*Shirashoola*)<br>follow up: range 07 days to 60 days <sup>a,b</sup> | 58 (3 RCTs) | ⊕□□□<br>VERY LOW <sup>d,f,g,h,i</sup> | - | The mean Nasal discharge ranged from - | SMD **0.43 lower**<br>(1.31 lower to |
|  |  |  |  | 2.22 to -1.2 | 0.46 higher) |
| Adverse Events (related to procedural Therapies) | No trial reported on outcome “Adverse Events (related to procedural Therapies)” |  |  |  |  |
| Other Adverse events | No trial reported on the outcome “Other Adverse events” |  |  |  |  |
| *The risk in the intervention group (and its 95% confidence interval) is based on the assumed risk in the comparison group and the **relative effect** of the intervention (and its 95% CI).<br>**CI:** Confidence interval; **SMD:** Standardised mean difference, **ST:** Single Therapy, **CT:** Combined Therapy, **PT:** Procedural therapy, **NPT:** Non-procedural therapy |  |  |  |  |  |
| **Explanations**<br>a. Assessed with five points Likert scale, 0 to 4, where 4 means worse, the scale was developed by authors but data about validated and reliability was not available.<br>b. Not mentioned in all included studies.<br>c. Each comparison in a multiple arm study was considered as an individual study.<br>d. Downgraded twice due to a high or unclear risk of bias across most domains and particularly around randomization, allocation concealment, and blinding with poor reporting of methods.<br>e. I<sup>2</sup> value for this analysis was <60%, did not downgraded for inconsistency.<br>f. As the PICOT of review question is broad so no further downgrade for indirectness.<br>g. Downgraded once for imprecision due to the very small sample size.<br>h. Not able to assess publication bias, because a small no. for studies(<10) were included in the analysis.<br>i. Downgraded one level for serious inconsistency (I<sup>2</sup> >60%) |  |  |  |  |  |

### Quality of the evidence

All the included trials were methodologically weak and poorly reported mostly in non-indexed journals, which results in the synthesis of evidence with ‘very low’ certainty in their results. There was insufficient or no information about randomization, allocation concealment, and blinding in most of the included trials which may result in a high chance of performance bias and detection bias[25]. The trials identified in this review were only Indian trails which compromise the generalizability of the results. We were unable to detect the publication bias as the number of included studies was insufficient for formal statistical testing and to generate funnel plots[10]. Although such limitations do not always mean that the treatment is not safe and ineffective, they might indicate that the effectiveness and safety have not been adequately investigated.

### Limitations

The question of this review was addressing a broad PICOT(Population, Intervention, Control, and Outcome, Timepoint), which included a broad range of conventional diagnostic sub-classification viz. Acute and chronic rhinosinusitis, maxillary sinusitis, as well as the various resembling similar conditions in Ayurveda e.g. *Peenasa, Dushta Pratishyaya, Kaphaja Shira Shoola, Kaphaja Shiroroga*, and *Suryavarta*. The criteria used to define disease conditions, the chronicity of conditions, and the age groups of participants were varied considerably among the included trials. About 13 complex Ayurveda formulations and 05 Ayurveda procedural interventions with different doses, duration, and schedules have been tested but none of these protocols have been tested repeatedly. The outcome measures were also varied among the trials and none of the trials included any quality of life(QoL) and safety outcome measures in their assessments. As we found only 6 trials for quantitative analysis, and the protocols of these trials were not accessible so the chance of publication bias and selective reporting bias cannot be ignored. Although we have highlighted the clinical diversity amongst the trials, it is important to understand whether there is a difference in the treatment effect between these two major treatment modalities of Ayurveda.

## CONCLUSION

### Implications for practice

*For clinicians:* Due to the lack of Ayurveda versus Placebo or Non-Ayurveda intervention trials, the authors are not able to make any conclusion. Although the results of individual studies comparing CT versus ST, appeared to favor CT, very low certainty of total effect(downgraded twice for RoB, once for inconsistency, indirectness, and imprecision each) hindered to arrive at any conclusion also in this category of comparison. *For funders of interventions:* It is highly recommended to conduct high-quality non-inferiority or equivalence efficacy and safety trials adopting an ‘Integrative Trial Design’ comparing Ayurveda interventions versus standard conventional care or Ayurveda as add on therapy versus conventional therapy, and Ayurveda versus placebo trials wherever justified.

### Implications for research

We found very poor adherence to reporting guidelines in most of the studies included in this review. It is recommended for authors, reviewers, and editors to comply with available reporting guidelines suitable for Ayurveda trials[26-28]. Also, the available guidelines[29], and tools[30] should also be utilized during protocol development. There is a need for the development of specific guidelines or extensions to existing guidelines for reporting Ayurveda trials.

Considering the limitation of blinding in some Ayurveda procedural intervention trials, blinding of outcomes assessors and allocation sequence concealment are needed to be implemented. The prospective meta-analysis methodology[31] may be adopted in conducting small trials as required for P.G. and Ph.D. courses.

There were around 50 different subjective and objective OMs assessed in studies included in this review, most of them were subjective and the information about how they were assessed was also missing and the validity and reliability of these criteria are also questionable. None of the trials included any safety assessment criteria for procedural as well as non-procedural interventions. There is a need to develop standard outcome measures and Core Outcome Sets for Ayurveda following standard development guidelines [32].

## Supporting information

Supplementary appendix_1_completed_PRISMA_checklist

Supplementary appendix_2_Review Protocol

Supplementary Appendix_3_Search strategy

Supplementary appendix_4_Contents of Interventions

Supplementary Appendix_5_Summary of Quantitative Analysis

## Data Availability

The datasets used and analyzed during the current study are available from the corresponding author on reasonable request.

## FUNDING FOR THIS REVIEW

This review was funded by Central Council for Research in Ayurvedic Sciences(CCRAS), Ministry of AYUSH, Government of India (Grant No. 3-13/2019-CCRAS/Admn/IMR/Systematic Review/Hq 1), and all authors are working as Research officers for this Council.

## ACKNOWLEDGMENTS

We would like to thank **Vd.Prof.K.S.Dhiman**, Director General, CCRAS for his guidance and support, and **Dr. Himanshu Negandhi**, Prof. IIPH, New Delhi for methodological inputs.

## AUTHOR CONTRIBUTIONS

AA and MG conceived the article. AA, MG, and PD drafted the protocol, evaluated the risk of bias, and contributed to GARDE analysis. AA and PD retrieve the literature, extracted, and analyzed the data. AA wrote this manuscript and SJ contributed to language editing. MG, BCS, SJ, SK, and SN gave suggestions for the discussions and structure of the manuscript. All authors have read and approved this manuscript.

## ETHICS APPROVAL AND CONSENT TO PARTICIPATE

Not applicable.

## CONSENT FOR PUBLICATION

Not applicable.

## SUPPORTING INFORMATION

S1 File. Review protocol

S2 File. Search strategy for AYUSH Research Portal and Cochrane CENTRAL databases.

S3 File. Interventions used in the included studies.

S4 File. Summary of Quantitative Analysis.

## REFERENCE

1. Fokkens WJ, Lund VJ, Mullol J, et al. EPOS 2012: European position paper on rhinosinusitis and nasal polyps 2012, A summary for otorhinolaryngologists. Rhinol Online. 2012 Mar;50(1):1–12.

2. The Royal College of Surgeons: Commissioning Guide-Rhinosinusitis 2016. https://www.rcseng.ac.uk/-/media/files/rcs/standards-and-research/commissioning/rhinosinusitis-commissioning-guide-for-republication.pdf (accessed 15 Oct 2020).

3. Pleis JR, Lucas JW, Ward BW. Summary health statistics for U.S. adults: National Health Interview Survey, 2008. Vital Health Stat 2009; 242: 1–157.

4. Rosenfeld RM, Piccirillo JF, Chandrasekhar SS, Brook I, et al. Clinical practice guideline (update): adult sinusitis. Otolaryngol Head Neck Surg. 2015 Apr;152(2 Suppl): S1–S39.

5. Dr.Ravindra Chandra Chaudhary. 6th Chapter: Pratishyaya. In: Dr. R.C. Chaudhary, eds. Illustrated Shalakya-Vigyana. Varanasi: Chaukhambha Orientalia 2005:44–59.

6. Ministry of AYUSH: Ayurveda standard treatment guidelines 2017. https://namayush.gov.in/content/standard-treatment-guidelines (accessed 15 Oct 2020).

7. Kenealy T, Arroll B. Antibiotics for the common cold and acute purulent rhinitis. Cochrane Database of Systematic Reviews 2013, Issue 6. Art. No.: CD000247. DOI:10.1002/14651858.CD000247.pub3. Accessed 09 April 2021.

8. Sridharan K, Mohan R, Ramaratnam S, Panneerselvam D. Ayurvedic treatments for diabetes mellitus. Cochrane Database of Systematic Reviews 2011, Issue 12. Art. No.: CD008288. DOI:10.1002/14651858.CD008288.pub2. Accessed 12 April 2021.

9. Agarwal V, Abhijnhan A, Raviraj P. Ayurvedic medicine for schizophrenia. Cochrane Database of Systematic Reviews 2007, Issue 4. Art. No.: CD006867. DOI:10.1002/14651858.CD006867. Accessed 12 April 2021.

10. Higgins JPT, Thomas J, Chandler J, et al. Cochrane Handbook for Systematic Reviews of Interventions version 6.1 (updated September 2020). Cochrane, 2020. https://training.cochrane.org/cochrane-handbook-systematic-reviews-interventions (accessed 15 Oct 2020).

11. Moher D, Liberati A, Tetzlaff J, et al. The PRISMA Group (2009) Preferred Reporting Items for Systematic Reviews and Meta-Analyses: The PRISMA Statement. PLOS Medicine 6(7): e1000097. doi:10.1371/journal.pmed.1000097.

12. Higgins JPT. Excel tool to implement RoB 2. https://sites.google.com/site/riskofbiastool/welcome/rob-2-0-tool/current-version-of-rob-2?authuser=0 (accessed 19 Sep 2020).

13. Schünemann H, Brożek J, Guyatt G, Oxman A, editors. GRADE handbook for grading quality of evidence and strength of recommendations. Updated October 2013. The GRADE Working Group, 2013. https://gdt.gradepro.org/app/handbook/handbook.html (accepted19 Sep 2020).

14. GRADEpro GDT: GRADEpro Guideline Development Tool [Software]. McMaster University, 2020 (developed by Evidence Prime, Inc.). https://gradepro.org/ (accessed 19 Sep 2020).

15. Review Manager (RevMan) [Computer program]. Version 5.3, The Cochrane Collaboration, 2020. https://training.cochrane.org/online-learning/core-software-cochrane-reviews/revman (accessed 19 Sep 2020).

16. Rajeev K Rail, K Govardhan, Ajay K Sharma. Management of Peenasa/Pratishyaya Roga (Rhinitis/Sinusitis) With Nasya Karma – A Clinical Study. Journal of Research in Ayurveda and Siddha. 2002;23:1–2:81-90.

17. Nagaraja JM. Role of Vidanga Taila Nasya in the management of Kaphaja Shira Shoola W.S.R. to Chronic Maxillary Sinusitis. M.D. Thesis, Rajiv Gandhi University of health sciences, Bangalore. 2009. http://52.172.27.147:8080/jspui/bitstream/123456789/5108/1/Nagaraja%20J%20M.pdf (accessed 20 Sep 2020).

18. Chaudhari V, Rajagopala M, Mistry S, et al. Role of Pradhamana Nasya and Trayodashanga Kwatha in the management of Dushta Pratishyaya with special reference to chronic sinusitis. AYU. 2010; doi:10.4103/0974-8520.77165.

19. Ashwini BN, K Siva Balaji. Role of Trikatu Taila Nasya in the Management of Kaphaja shirashoola w.s.r. to Maxillary Sinusitis. International Ayurvedic Medical Journal. 2015;3:1.

20. Parth P Dave, Kunjal H Bhatta, Vaghela DB, et al. Role of Vyaghri Haritaki Avaleha and Anu Taila Nasya in the management of Dushta Pratishyaya (Chronic Sinusitis). IJAM 2016;7:1.

21. Sarmah Jayanta, Mangal Gopesh. A clinical study to evaluate the efficacy of Vamana and Nasya Karma in the management of Peenasa w.s.r. to Sinusitis. Journal of Ayurveda. 2017;3:66–73.

22. Mansi Sharma A. A clinical study on the efficacy of Ardhanarishvara Rasa Nasya and Nimbadi Guggulu in the management of Kaphaja Shiroroga w.s.r. to sinusitis, Journal of Ayurveda. 2018;4:66–72.

23. Sanjay Sukhdeorao Thoka, DS Mishra, MK Shandilya. Comparative study of Vamana &Nasya Karma in the management of Peenasa Roga (Sinusitis). Journal of Research in Ayurvedic Sciences. 2008;29-2:29–38.

24. Sree Kumar K. Role of Anutaila Nasya and Varunadi Ghruita in the management of Dushta Pratishyaya w.s.r. to chronic sinusitis. M.D. Thesis, Gujarat Ayurveda University, Jamnagar, Ayurveda Research Database-ARD. 2005 https://ayurvedahealthcare.info/content/ayurveda-research-database-ard. (accessed 20 Sep 2020).

25. Schulz KF, Chalmers I, Hayes RJ, et al. Dimensions of methodological quality associated with estimates of treatment effects in controlled trials. JAMA 1995;273(5):408–12.

26. Gagnier JJ, Boon H, Rochon P, et al. CONSORT Group. Reporting randomized, controlled trials of herbal interventions: an elaborated CONSORT statement. Ann Intern Med. 2006 Mar 7;144(5):364–7. doi:10.7326/0003-4819-144-5-200603070-00013.

27. Boutron I, Altman DG, Moher D et al. CONSORT NPT Group. CONSORT Statement for Randomized Trials of Nonpharmacologic Treatments: A 2017 Update and a CONSORT Extension for Nonpharmacologic Trial Abstracts. Ann Intern Med. 2017 Jul 4;167(1):40–47. doi:10.7326/M17-0046.

28. Cheng CW, Wu TX, Shang HC, et al. CONSORT-CHM Formulas 2017 Group. CONSORT Extension for Chinese Herbal Medicine Formulas 2017: Recommendations, Explanation, and Elaboration. Ann Intern Med. 2017 Jun 27;167(2):112–121. doi:10.7326/M16-2977.

29. Chan A-W, Tetzlaff JM, Altman DG, et al. SPIRIT 2013 Statement: Defining standard protocol items for clinical trials. Ann Intern Med. 2013;158(3):200–207.

30. Loudon Kirsty, Treweek Shaun, Sullivan Frank, et al. The PRECIS-2 tool: designing trials that are fit for purpose. BMJ. 2015. doi:10.1136/bmj.h2147.

31. Seidler Anna Lene, Hunter Kylie E, Cheyne Saskia, et al. A guide to prospective meta-analysis. BMJ. 2019. doi:10.1136/bmj.l5342.

32. Kirkham JJ, Gorst S, Altman DG, et al. Core Outcome Set–STAndards for Reporting: The COS-STAR Statement. PLoS Med. 2016. doi:10.1371/journa.

