## Supplementary appendix_1_completed_PRISMA_checklist for "Efficacy and safety of Ayurveda interventions for Sinusitis: A systematic review and meta-analysis"

### Citation

AZEEM AHMAD, MANOHAR S. GUNDETI, PARTH P. DAVE. Efficacy and safety of ayurveda interventions for sinusitis: a systematic review. PROSPERO 2018 CRD42018103995 Available from:
https://www.crd.york.ac.uk/prospero/display_record.php?ID=CRD42018103995

### Review question

1.What is the efficacy and safety of Ayurveda interventions for the management of Sinusitis.

2.What is the relative efficacy and safety of *Shodhana* and *Shamana* Ayurveda treatment modalities for the management of Sinusitis.

### Searches

Following database will be search-
PubMed
Cochrane Librery(Cochrane Central Register of Controlled Trials : Issue 6 of 12, June 2018)
AYUSH Research Portal (Govt of India)
DHARA
Google Scholar
Online clinical trials registers

### Types of study to be included

Randomized controlled trials (RCTs), quasi-randomized controlled trials, controlled clinical trials (CCTs), multiple arms clinical trial that are of at least 3 weeks duration will be eligible for inclusion.

### Condition or domain being studied

Due to the increased environmental pollution, faulty lifestyle and decreased immunity; rhinitis (acute nonspecific rhinitis- common cold) is one of the most common acute infections affecting the body. inadequately treated rhinitis will cause the spread of infection into sinuses and results into sinusitis.
The prevalence of sinusitis (146/1000 population) has been reported to exceed that of any other chronic condition.
The goal of treatment is to re-establish sinus ventilation and to correct mucosal opposition and stop the spread of infection.
The features of the disease *Pratishyaya* mentioned in different Ayurveda texts is similar to that of sinusitis/ rhinosinustis in modern science. Many treatment modalities propounded by ancient scholars for this disease according to condition of patient and progression of disease. Two main types of treatment modalities in Ayurveda are *Shodhana* (purification) and *Shamana* (pacification).*Nasya* (nasal instillation of medicine) is one of the choice of the treatment modality for sinusitis in Ayurveda. Also there are many *shamana* drugs are mentioned in Ayurveda text but very few of them have been tested clinically in present era. Along with efficacy, safety of these modalities and quality of these trials should also be analysed and critically evaluated and make it highlighted in public domain.

#### Participants/population

Patients fulfilling the diagnostic criteria based on symptomatology of Pratishyaya/Pinasa explained in Ayurvedic classics and sinusitis irrespective of age and sex will be selected.

Subjective Criteria- Patients having symptoms of nasal discharge, nasal blockage, local pain, headache, nasal stuffiness, anosmia etc.

Objective criteria- Radiological examination of paranasal sinuses and hematological examination.

Exclusion- Patients who do not meet the diagnostic criteria or cases which require surgical treatment.

#### Intervention(s), exposure(s)

Ayurveda Treatment (Shamana or/and Shodhana) with any dose, type, schedule, medicine, medicine form and Pathayapathya.

(Patients may receive additional non-Ayurveda intervention in all groups of study)

#### Comparator(s)/control

1. Ayurveda Treatment (Shamana or/and Shodhana) with different dose, type, schedule, medicine, medicine form as compare to intervention(s)/ exposure(s).

2. Placebo and/or Sham therapy

3. Non-Ayurveda interventions or combination of Ayurveda and non Ayurveda interventions.

#### Context

No restriction

#### Main outcome(s)

1.Response to treatment (improvement in Subjective and/or objective criteria of assessment)

2.Serious adverse events(resulting in death, disability or incapacity, complications, were life threatening, led to hospitalization or prolong of hospitalization

***Measures of effect***

Administration timings vary from 7 days to 45 days as different categories of medications are included for review.

#### Additional outcome(s)

1.Withdrawals due to adverse events or lack of efficacy or inconvenience of therapy/treatment.

2.Number of patients with specific adverse event.

***Measures of effect***

During the study period or up to one month after completion study.

#### Data extraction (selection and coding)

Two review authors Dr. Azeem Ahmad & Dr. Parth Dave independently will assess the title and abstract identified during the search. Potentially eligible articles were read in full to determine whether they meet the eligibility criteria. Disagreements were discussed with third review author Dr. Manohar Gundeti. If necessary,

additional information will be obtained from the contact person (authors) of that study through e-mail or telephone.
We will make a predesigned form to extract data from the included studies for assessment of study quality and data analysis. Data will be extracted independently by two review authors Dr. Azeem Ahmad & Dr. Parth Dave. Any disagreement will be consulted and settled through discussions with a third author Dr. Manohar Gundeti, where necessary.
All three review authors will assess the quality of reporting trial independently as follows:
1. CONSORT (Consolidated Standards of Reporting Trials)-2010 check list: for quality assessment of included parallel group randomized clinical trial.
2. TREND (Transparent Reporting of Evaluations with Non-randomized Designs)- 2004 check list for quality assessment of included non-randomized trial.
Assessment will be done under three categories, 'Yes' reporting, 'No' reporting and 'Incomplete' reporting. 2 points will be given for each item if it is reported completely, in case of incomplete reporting only one point will be given to that item and no point for 'No' reporting. Results will be interpreted in terms of Percentage (%) of mean of each three category reporting items.

#### Risk of bias (quality) assessment

Two authors (Dr. Azeem Ahmad and Dr. Parth Dave) will independently assess the risk of bias in included studies.
RCTs will be assessed with the help of Cochrane tool of Risk of bias and that of non-randomized trials will be assessed with ROBINS-I tool (Risk Of Bias In Non-randomized Studies - of Interventions).
Disagreements between these review authors will be resolved by discussion, with involvement of a third review author (MG) if necessary and the results will be interpreted.

#### Strategy for data synthesis

We will narratively synthesize the results and present the results in count, percentage and frequency.

#### Analysis of subgroups or subsets

NA

#### Contact details for further information

Azeem Ahmad


#### Organisational affiliation of the review

CENTRAL COUNCIL FOR RESEARCH IN AYURVEDIC SCIENCES
http://www.ccras.nic.in/

#### Review team members and their organisational affiliations

Dr AZEEM AHMAD. CENTRAL COUNCIL FOR RESEARCH IN AYURVEDIC SCIENCES
Dr MANOHAR S. GUNDETI. CENTRAL COUNCIL FOR RESEARCH IN AYURVEDIC SCIENCES
Dr PARTH P. DAVE. CENTRAL COUNCIL FOR RESEARCH IN AYURVEDIC SCIENCES

#### Collaborators

Dr Saketh Ram Thrigulla. National Institute of Indian Medical Heritage, Hyderabad

#### Type and method of review

Intervention, Narrative synthesis, Systematic review

#### Anticipated or actual start date

05 July 2018

**Anticipated completion date**

15 September 2018

**Funding sources/sponsors**

CCRAS, Ministry of AYUSH, Government of India

**Conflicts of interest**

**Language**

English

**Country**

India

**Stage of review**

Review Completed not published

**Subject index terms status**

Subject indexing assigned by CRD

**Subject index terms**

Humans; Medicine, Ayurvedic; Sinusitis

**Date of registration in PROSPERO**

31 August 2018

**Date of first submission**

15 August 2018

**Details of any existing review of the same topic by the same authors**

**Stage of review at time of this submission**

| Stage | Started | Completed |
|---|---|---|
| Preliminary searches | Yes | Yes |
| Piloting of the study selection process | Yes | Yes |
| Formal screening of search results against eligibility criteria | Yes | Yes |
| Data extraction | Yes | Yes |
| Risk of bias (quality) assessment | Yes | Yes |
| Data analysis | Yes | Yes |

**Revision note**

Review is completed

*The record owner confirms that the information they have supplied for this submission is accurate and complete and they understand that deliberate provision of inaccurate information or omission of data may be construed as scientific misconduct.*

*The record owner confirms that they will update the status of the review when it is completed and will add publication details in due course.*

#### Versions

31 August 2018
22 February 2019
24 July 2019

PROSPERO
This information has been provided by the named contact for this review. CRD has accepted this information in good faith and registered the review in PROSPERO. The registrant confirms that the information supplied for this submission is accurate and complete. CRD bears no responsibility or liability for the content of this registration record, any associated files or external websites.
