## Supplementary appendix_2_Review Protocol for "Efficacy and safety of Ayurveda interventions for Sinusitis: A systematic review and meta-analysis"

**Supplementary Appendix 1.**

| **Search strategy** | |
| --- | --- |
| **For AYUSH Research Portal** | HOME>AYURVEDA>CLINICAL RESEARCH>RESPIRATORY>SINUSITIS ACUTE/CHRONIC (ICPC-R75/ICD-J01,J32)  SEARCH >Ayurveda>Sinusitis  SEARCH >Ayurveda>pinasa |
| **Cochrane CENTRAL** | http://cochranelibrary-wiley.com/cochranelibrary/search/ SINUSITIS AND AYURVED* (under the title, abstract, keywords) PINASA AND AYURVED* (under the title, abstract, keywords) |
