## Supplementary Appendix_3_Search strategy for "Efficacy and safety of Ayurveda interventions for Sinusitis: A systematic review and meta-analysis"

| **Type of Therapy*** | | **Medicine Used** | **Dosage Used** | **Duration (Days)** | **Contents** |
| --- | --- | --- | --- | --- | --- |
| **Procedural(*Shodhana)* therapies(PT)** | ***Nasya Karma*** | *Pathadi Taila^[1]^* | Not Mentioned | 14-21 | Cissampelos pareira *Linn,* Curcuma longa *L,* Berberisaristata *DC,* Sansevieria roxburghiana*. Schult. &Schult.f.,* Piper longum *Linn,* Jasminum grandiflorum *Linn.,* Baliospermum montanum *(Willd.) Muell.-Arg.,* Seasom indicum *L.* |
|  |  | *Anutaila^[2]^* | 4-8 DsPN/D | 7-21 | Leptadenia reticulata *(Retz.) Wight &Arn,* Coleus vettiveroides *K.C. Jacob,* Cedrus deodara *- (Roxb. ex D.Don.)G.Don.,* Cyperus rotundus *L.,* Cinnamomum zeylanica *Blume,* Vetiveria zizanoides *Linn.,* Hemidesmus indicus *(L.) R. Br. ex Schult.,* Berberis aristata *DC,* Glycyrrhiz aglabra *Linn,* Cyperus platystylis *R.Br.,* Aquilaria agallocha *Roxb.,* Asparagus racemosus *(Willd.),* Saccharum officinarum *L.,* Aegle marmelos *L.,* Nymphaea stellata *Willd.,* Solanum indicum *Linn.,* Solanum xanthocarpum *Schrad. &Wendl. ,* Boswellia serrata *Triana & Planch.,* Uraria picta *Desv.,* Desmodium gangetium *DC Pennel.,* Embelia ribes *Burm F.,* Cinnamomum tamala*(Buch. -Ham.) Nees&Eberm,.* Elettaria cardamomum *Maton,* Vitex negundo *L.,* Nelumbo nucifera *Gaertn. ,* Abutilon indicum *L.,* Sesamum indicum *L. ,* and Aqua pluvialis*.* |
|  |  | *Trikatu Taila* | 8 DsPN/D | 7 | Piper nigrum *Linn.,* Piper longum *Linn.,* Zingiber officinalale *Roscoe, and* Seasom indicum *L.* |
|  |  | *Sarsapa Taila* | 8 DsPN/D | 7 | Brassica campestris *Linn.* |
|  |  | *Vyaghri Taila^[3]^* | 8 DsPN/D | 7 | *Solanum surrattense, Baliospermum montanum Muel-Arg., Acorus calamus Linn., Moringa olifera Linn., Ocimum sanctum Linn., Zingiber officinalale Roscoe, Piper nigrum Linn., Piper longum Linn., Rock salt., Seasom indicum L. and* Water*.* |
|  |  | *Vidanga Taila^[4]^* | 8 DsPN/D | 7 | *Embelia ribes Burm.f., Seasom indicum L.* |
|  |  | *Tila Taila* | 8 DsPN/D | 7 | *Seasom indicum L.* |
|  |  | *Brahmi Ghruta^[5]^* | 6 DsPN/D | 14 | *Bacopa monnieri (L.)Pennel, Acorus calamus Linn., Saussurea lappa C.B.Clarke, Convolvulus pluricaulis Choisy,* and cow’s ghee.^[6]^ |
|  |  | *Ardhanarishvara Rasa^[7]^* | 4 DsPN/D | 14 | Varatikabhasma(fine powder of external shell of sea animal Cypreamonetalinn.), Tankana(Borax fine powder), *Piper nigrum Linn.,* *Aconitum ferox*, Cow’s Milk. |
|  |  | *Trikatu and Triphal* powder | 2.5–7.5 gm/nostril | 7 | *Piper nigrum Linn., Piper longum Linn., Zingiber officinalale Roscoe, and Terminalia chebula Retz., Terminalia bellerica Roxb., Emblica officinalis Gaertn.* |
|  | ***Snehapana*** | *GuggulutiktakGhruta^[8]^* | Increasing dose pattern (details not mentioned) | 7 | *Azadirachtaindica A. Juss, Trichosanthes dioica Roxb., Solanum xanthocarpum Schrad and Wendl., Tinospora cordifolia (Thunb.) Miers, Adhatoda vasica Nees., purified Commiphoramukul (Hook. Ex Stocks), Cissampelos pareira L., Embelia ribes Burm F., Cedrus deodara (Roxb. ex D.Don.) G.Don, Piper chaba Hunter, Hordeum vulgare L., Zingiber officinale Rosc., Curcuma longa L., Anethum sowa L., Saussurea lappa Clarke, Zanthoxylum alatum Roxb., Piper nigrum Linn., Holarrhena antidysenterica (L.) Wall. ex A. DC., Trachyspermum ammi L. Sprague, Plumbago zeylanica L, Picrorhiza kurroa Royle ex Benth,* purified *Semecarpus anacardium Linn., Acorus calamus. (AC) Linn., Piper longum Linn., Pluchea lanceolata (DC.) Oliv. &Hiern, Rubia cordifolia L., Aconitum heterophyllum Wallich., Aconitum ferox Pennel., Trachyspermum copticum Linn.* |
|  | ***Vamana*** | Not Mentioned | One sitting | NA | Not Mentioned |
|  | ***Virechana*** | *Trivritlehya^[9]^* | One sitting | NA | *Saccharum officinarum L.,* Merremia turpethum *(L.) Rendle, Elettaria cardamomum (L.) Maton,*Cinnamomum verum*. J.Presl, Cinnamomum tamala · (Buch. -Ham.) Nees&Eberm, Honey* |
|  | ***NadiSwedana*** | *DashamulaKwatha^[10]^* | 15 min, BD | 7-21 | *Gmelina arborea* Roxb., *Aegle marmelos* (Linn.) Correa, *Stereospermum colais* (Dillw.) Mabb., *Oroxylum indicum* Vent., *Premna corymbosa* Rottler&Willd., *Pseudarthria viscida* (L) Wight and Arn., *Desmodium gangeticum* (L.) DC, *Solanum anguivi Lam*., *Solanum virginianum* L., *Tribulusterrestris* L. |
| ***Non-Procedural(Shamana)* Therapies(NPT)** | ***Kvatha/ Sheeta/ Fanta*** | *PatyakshadhatriPhanta^[11]^* | BD | 28 | *Terminalia chebula Retz., Terminalia bellerica Roxb., Emblica officinalis Gaertn., Andrographis paniculata (Burm. F), Curcuma longa L., Azadirachta indica A. Juss, Tinospora cordifolia (Thunb.) Miers.* |
|  |  | *TrayodashangaKwatha^[12]^* | 45 ml, BD | 45 | *Gmelina arborea* Roxb., *Aegle marmelos* (Linn.) Correa, *Stereospermum colais* (Dillw.) Mabb., *Oroxylum indicum* Vent., *Premna corymbosa* Rottler&Willd., *Pseudarthria viscida* (L) Wight and Arn., *Desmodium gangeticum* (L.) DC, Solanum anguivi Lam., *Solanum virginianum* L., *Tribulus terrestris* L., *Uraria picta Desv., Desmodium gangetium DC Pennel., Piper longum Linn., Zingiber officinalale Roscoe,Coriandrum sativum Linn.* |
|  | ***Vati*** | *NimbadiGuggulu^[13]^* | 2 tbs (500 mg)BD | 30 | *Azadirachtaindica A. Juss, Terminalia chebula Retz., Terminalia bellericaRoxb., EmblicaofficinalisGaertn., TrichosanthesdioicaRoxb., Adhatodavasica (L.) Nees, Commiphoramukul (Hook. Ex Stocks),* |
|  | ***Avleha*** | *VyaghriHaritaki Avaleha^[14]^* | 5 to 10 gm, BD | 6 | *Solanum xanthocarpum Schrad and Wendl.*, *Terminalia chebula Retz.*,*Zingiber officinalale Roscoe*, *Piper nigrum Linn., Piper longum Linn., Cinnamomum zeylanica Blume*, *Cinnamomum tamala (Buch. -Ham.) Nees&Eberm.,* *Elettaria cardamomum (L.) Maton, Mesuaferrea Linn., saccharum officinarum* |
|  | ***Ghrita*** | *Varunadi Ghrita^[15]^* | 10 gms, BD | 30 | *Crataeva magna, Barleria strigose*, *Asparagus racemosus, Plumbago zeylanica, Chonemorpha fragrans, Aegle marmelos, Gymnema sylvestre, Solanum anguivi, Solanum virginianum, Pongamia pinnata, Holoptelea integrifolia, Premna corymbose, Terminalia chebula, Moringa concanensis, Desmostachya bipinnata, Semecarpus anacardium, Crataeva magna, Barleria strigose, Asparagus racemosus, Plumbago zeylanica, Chonemorpha fragrans, Aegle marmelos, Gymnema sylvestre, Solanum anguivi, Solanum virginianum, Pongamia pinnata, Holoptelea integrifolia, Premna corymbose, Terminalia chebula, Moringa concanensis, Desmostachya bipinnata, Semecarpus anacardium.* |
|  | ***Rasa-aushadhi*** | *Shringarabhra Rasa^[16]^* | 250 mg, BD | 21 | *Cinnamomum camphora (L.) Nees&Eberm., Pavonia odorata Willd.,* Myristica fragrans *Houtt.,* Piper chaba *Hunter,* Cinnamomum tamala *(Buch. -Ham.) Nees&Eberm.,* Cinnamomum zeylanicum *Blume,* Abies webbiana *Lindl.,* Nardostachys jatamansi *DC.,* Syzygium zeylanicum *(L.) DC.,* Mesua ferrea *Linn.,* Saussurea lappa *C.B.Clarke,* Woodfordia fruticosa *Kurz,* Piper nigrum *Linn.,* Piper longum *Linn.,* Zingiber officinalale *Roscoe,* Terminalia chebula *Retz.,* Terminalia bellerica *Roxb.,* Emblicaofficinalis *Gaertn.,* Sulphurium *(Sulphar),* Hydrargyrum *(Mercury),* Calx of mica. |
| ***Definitions & Standard Ayurveda Terminology Codes^[17]^:**  ***Shodhana-*** major purification therapy/ bio-cleansing therapy/ detoxification therapy( SAT-I.76).  ***Shamana-***pacification of vitiated doṣa( SAT-F.29)  ***Nasya Karma***- medication through nasal route(SAT-I.156);  ***Snehapana***-therapeutic intake of medicated unctuous substance( SAT-I.445);  ***Vamana***-therapeutic process for controlled induction of vomiting along with a series of pre and post-operative measures.( SAT-I.139);  ***Virechana***- therapeutic process for controlled induction of purgation along with a series of pre and post-operative measures( SAT-I.140);  ***Nadi Swedana***- sudation using pipe like instrument( SAT-I.112)  ***Kvatha-*** decoction obtained by boiling coarse powder of drug(s) in specified proportions of water and reduced to a certain amount( SAT-G.42)  ***Phanata***- the infusion obtained by pouring boiling water on the powdered drug(s) and used after filtering( SAT-G.45)  ***Vati-***Pills  ***Churna-***fine sieved powder of well-dried drug(s)( SAT-G.43)  ***Avleha-*** Medicated jam  ***Ghrita-***Medicated Clarified Butter(Ghee)  ***Rasa-aushadhi-*** Metallo-mineral preparations  **Abbreviation**  **DsPN/D**: Drops per Nostril/Day; PT: Procedural Therapy; NPT: Non-Procedural Therapy | | | | | |

**References**

1. Acharya Agnivesa. Trimarmiyachikitsaadhyaya of Chikitsasthan, Verse 145. In: Achrya Vidyadhar Shukla, Prof. Ravidutta Tripathi, editors. Charak Samhita Volume-II with Vaidyamanorama hindi commentary. Varanasi: Chaukhambha Sanskrit Sansthan; 2012. p. 648.
2. Acharya Vagabhatta. Nsyavidhiaadhyaya of Sutrasthana. In: Vaidya Yadunandan Upadhyaya, editor. Astanga Hridaya. Varanasi: Chaukhambha Sanskrit Sansthan; 2003. p. 127.
3. Chakrapanidatta. Nasa Roga Adhikara, verse 5. In: Indradeva Tripathi, editor. Chakradatta. Varansi: Chaukhambha Sanskrit series; 1997, p. 343.
4. Acharya Vagabhatta. Shirorogapratisedhaadhyaya, Verse 7. In: Atridevavidhyaalankar, editor. Astanga Sangraha. Varanasi: BHU press; 1962. p. 289.
5. Bhavamishra. Apasmarachikitsaadhyaya, verse 18. In: Dr. Bulusu Sitaram, editor. Bhavaprakasha of Bhavamishra Vol-II. Varanasi: ChoukhambaSamskrutaSansthana; 2005. p. 225.
6. Anonymous. The Indian Pharmacopoeia. Vol. 2. New Delhi: Govt of India publication; 1996.
7. Siddhi Nandan Mishra. Chapter 65, verse 48-49. In: Govind Das, editor. Bhaishajya Ratnavali. Varanasi: Chaukhambha Surbharati Prakashan; 2013, p.1017
8. Acharya Vagabhatta. Vatavyadhichikitsaadhyaya of Chikitsasthana, verse 57-60. In: Vaidya Yadunandan Upadhyaya, editor. Astanga Hridaya. Varanasi: Chaukhambha Sanskrit Sansthan; 2003. p. 420.
9. Acharya Vagabhatta. VIrechankalpaadhyaya of Kalpasthana, verse 9. In: Vaidya Yadunandan Upadhyaya, editor. Astanga Hridaya. Varanasi: Chaukhambha Sanskrit Sansthan; 2003. p. 433.
10. Das G. Bhishagratna. Kasarogadhikara. In: Shastri R, Mishra BV, Shastri AD, editors. BhaishajyaRatnavali Hindi Commentary Analysis with Appendixes. Varanasi: Chaukhambha Sanskrit Sansthan; 2005. p. 13-15.
11. Acharya Sharangdhara. Madhyamakhanda, Qwatha Kalpana, verse 143-145. In: Prof. K.R.Srikanta Murthy, editor. Sharangdhara Samhita. Varanasi: Chaukhambah Orientalia; 1995. p. 73.
12. Das G. Bhishagratna. Rajayakshma Nidana. In: Shastri R, Mishra BV, Shastri AD, editors. BhaishajyaRatnavali Hindi Commetary Analysis with Appendixes. Varanasi: Chaukhambha Sanskrit Sansthan; 2005. p. 21.
13. Dattram Sh. Krishanlal Mathur. In: Khemraja, editor. Brihat Nighantu Ratnakar Part 6. Mumbai: Khemraja Shri Krishandas Prakashan; 1981. P. 398.
14. Das G. Bhishagratna. Kasarogadhikara. In: Shastri R, Mishra BV, Shastri AD, editors. BhaishajyaRatnavali Hindi Commentary Analysis with Appendixes. Varanasi: Chaukhambha Sanskrit Sansthan; 2005. p. 43-46
15. Acharya Vagabhatta. Shodhanadigansangrah aadhyaya of Sutrasthana. In: Vaidya Yadunandan Upadhyaya, editor. Astanga Hridaya. Varanasi: Chaukhambha Sanskrit Sansthan; 2003. p. 104-108.
16. Das G. Bhishagratna. RajyakshmaRogadhikara In: Shastri R, Mishra BV, Shastri AD, editors. BhaishajyaRatnavali Hindi Commentary Analysis with Appendixes. Varanasi: Chaukhambha Sanskrit Sansthan; 2005. p. 212-221.
17. NAMASTE Portal. National AYUSH Morbidity and Standard terminology Electronic Portal; <http://www.namstp.ayush.gov.in/#/sat>. Accessed on 24.11.2020.
