## Supplementary appendix_4_Contents of Interventions for "Efficacy and safety of Ayurveda interventions for Sinusitis: A systematic review and meta-analysis"

**Supplementary Appendix 3**

| **Summary of Quantitative Analysis** | | | | |
| --- | --- | --- | --- | --- |
| **Outcome Measures & Comparisons** | **Comparisons (participants)** | **Effect Size SMD/RR [CI, 95%]** | **P-value (*I^2^)*** | **Included Studies(Ref.)** |
| **Nasal discharge** |  |  |  |  |
| CT Vs ST | 10(210) | -0.71 [-1.16, -0.26] | p=0.002(58%) | Rajeev et al., Sanjay et.al., Shreekumar et.al, Parth et al., Mansi et.al., Varsha et al. |
| CT Vs NPT | 7(149) | -0.89 [-1.53, -0.25] | p=0.007(69%) | Rajeev et al., Sanjay et.al., Shreekumar et.al, Parth et al., Mansi et.al., Varsha et al. |
| CT Vs PT | 3(61) | -0.38 [-0.88, 0.13] | p=0.15(0%) | Rajeev et al., Shreekumar et.al, Varsha et al. |
| PT Vs NPT | 3(58) | -0.41 [-1.22, 0.40] | p=0.32(56%) | Rajeev et al, Shreekumar et.al, Varsha et al. |
| **Nasal obstruction** |  |  |  |  |
| CT Vs ST | 10(216) | -0.31 [-0.74, 0.12] | p=0.16(57%) | Rajeev et al., Sanjay et.al., Shreekumar et.al, Parth et al., Mansi et.al., Varsha et al. |
| CT Vs NPT | 7(158) | -0.22 [-0.78, 0.33] | p=0.43(65%) | Rajeev et al., Sanjay et.al., Shreekumar et.al, Parth et al., Mansi et.al., Varsha et al. |
| CT Vs PT | 3(58) | -0.52 [-1.16, 0.13] | p=0.03(31%) | Rajeev et al, Shreekumar et.al, Varsha et al. |
| PT Vs NPT | 3(58) | -0.26 [-2.01, 1.48] | p=0.77(89%) | Rajeev et al, Shreekumar et.al, Varsha et al. |
| **Loss of smell** |  |  |  |  |
| CT Vs ST | 6(88) | -0.29 [-0.57, -0.02] | p=0.03(0%) | Shreekumar et.al, Parth et al., Varsha et al., Rajeev et al. |
| CT Vs NPT | 3(40) | -0.22 [-0.63, 0.19] | p=0.30(0%) | Shreekumar et.al, Parth et al., Varsha et al. |
| CT Vs PT | 3(48) | -0.35 [-0.71, 0.01] | p=0.06(13%) | Shreekumar et.al, Varsha et al., Rajeev et al. |
| PT Vs NPT | 2(28) | -0.16 [-1.46, 1.14] | p=0.81(64%) | Shreekumar et.al, Varsha et al. |
| **Headache** |  |  |  |  |
| CT Vs ST | 10(218) | -0.44 [-0.86, -0.02] | p=0.04(56%) | Rajeev et al., Sanjay et.al., Shreekumar et.al, Parth et al., Mansi et.al., Varsha et al. |
| CT Vs NPT | 7(157) | -0.55 [-1.06, -0.05] | p=0.03(57%) | Rajeev et al., Sanjay et.al., Shreekumar et.al, Parth et al., Mansi et.al., Varsha et al. |
| CT Vs PT | 3(61) | -0.16 [-1.02, 0.71] | p=0.72(64%) | Rajeev et al, Shreekumar et.al, Varsha et al. |
| PT Vs NPT | 3(58) | -0.43 [-1.31, 0.46] | p=0.34(63%) | Rajeev et al, Shreekumar et.al, Varsha et al. |
| **Abbreviations:**  Ay: Ayurveda; CT: Combined Therapy; ST: Single Therapy; PT: Procedural Therapy; NPT; Non-Procedural Therapy; SMD: Standard Mean Difference; RR: Risk Ratio; CI: Confidence Interval; NA: Not Applicable | | | | |
