## Supplementary Appendix_5_Summary of Quantitative Analysis for "Efficacy and safety of Ayurveda interventions for Sinusitis: A systematic review and meta-analysis"

### Reporting checklist for systematic review and meta-analysis.

Based on the PRISMA guidelines.

#### Instructions to authors

Complete this checklist by entering the page numbers from your manuscript where readers will find each of the items listed below.

Your article may not currently address all the items on the checklist. Please modify your text to include the missing information. If you are certain that an item does not apply, please write "n/a" and provide a short explanation.

Upload your completed checklist as an extra file when you submit to a journal.

In your methods section, say that you used the PRISMAreporting guidelines, and cite them as:

Moher D, Liberati A, Tetzlaff J, Altman DG, The PRISMA Group. Preferred Reporting Items for Systematic Reviews and Meta-Analyses: The PRISMA Statement

|  |  | Reporting Item | Page Number |
| --- | --- | --- | --- |
| **Title** |  |  |  |
|  | [#1](https://www.goodreports.org/reporting-checklists/prisma/info/#1) | Identify the report as a systematic review, meta-analysis, or both. | 1 |
| **Abstract** |  |  |  |
| Structured summary | [#2](https://www.goodreports.org/reporting-checklists/prisma/info/#2) | Provide a structured summary including, as applicable: background; objectives; data sources; study eligibility criteria, participants, and interventions; study appraisal and synthesis methods; results; limitations; conclusions and implications of key findings; systematic review registration number | 2,3 |
| **Introduction** |  |  |  |
| Rationale | [#3](https://www.goodreports.org/reporting-checklists/prisma/info/#3) | Describe the rationale for the review in the context of what is already known. | 3 |
| Objectives | [#4](https://www.goodreports.org/reporting-checklists/prisma/info/#4) | Provide an explicit statement of questions being addressed with reference to participants, interventions, comparisons, outcomes, and study design (PICOS). | 4 |
| **Methods** |  |  |  |
| Protocol and registration | [#5](https://www.goodreports.org/reporting-checklists/prisma/info/#5) | Indicate if a review protocol exists, if and where it can be accessed (e.g., Web address) and, if available, provide registration information including the registration number. | 4 |
| Eligibility criteria | [#6](https://www.goodreports.org/reporting-checklists/prisma/info/#6) | Specify study characteristics (e.g., PICOS, length of follow-up) and report characteristics (e.g., years considered, language, publication status) used as criteria for eligibility, giving rational | 4,2 |
| Information sources | [#7](https://www.goodreports.org/reporting-checklists/prisma/info/#7) | Describe all information sources in the search (e.g., databases with dates of coverage, contact with study authors to identify additional studies) and date last searched. | 5 |
| Search | [#8](https://www.goodreports.org/reporting-checklists/prisma/info/#8) | Present full electronic search strategy for at least one database, including any limits used, such that it could be repeated. | 5 |
| Study selection | [#9](https://www.goodreports.org/reporting-checklists/prisma/info/#9) | State the process for selecting studies (i.e., for screening, for determining eligibility, for inclusion in the systematic review, and, if applicable, for inclusion in the meta-analysis). | 5 |
| Data collection process | [#10](https://www.goodreports.org/reporting-checklists/prisma/info/#10) | Describe the method of data extraction from reports (e.g., piloted forms, independently by two reviewers) and any processes for obtaining and confirming data from investigators. | 5 |
| Data items | [#11](https://www.goodreports.org/reporting-checklists/prisma/info/#11) | List and define all variables for which data were sought (e.g., PICOS, funding sources), and any assumptions and simplifications made. | 5 |
| Risk of bias in individual studies | [#12](https://www.goodreports.org/reporting-checklists/prisma/info/#12) | Describe methods used for assessing risk of bias in individual studies (including specification of whether this was done at the study or outcome level, or both), and how this information is to be used in any data synthesis. | 6 |
| Summary measures | [#13](https://www.goodreports.org/reporting-checklists/prisma/info/#13) | State the principal summary measures (e.g., risk ratio, difference in means). | 6 |
| Planned methods of analyis | [#14](https://www.goodreports.org/reporting-checklists/prisma/info/#14) | Describe the methods of handling data and combining results of studies, if done, including measures of consistency (e.g., I2) for each meta-analysis. | 6 |
| Risk of bias across studies | [#15](https://www.goodreports.org/reporting-checklists/prisma/info/#15) | Specify any assessment of risk of bias that may affect the cumulative evidence (e.g., publication bias, selective reporting within studies). | 6 |
| Additional analyses | [#16](https://www.goodreports.org/reporting-checklists/prisma/info/#16) | Describe methods of additional analyses (e.g., sensitivity or subgroup analyses, meta-regression), if done, indicating which were pre-specified. | 6 |
| **Results** |  |  |  |
| Study selection | [#17](https://www.goodreports.org/reporting-checklists/prisma/info/#17) | Give numbers of studies screened, assessed for eligibility, and included in the review, with reasons for exclusions at each stage, ideally with a [flow diagram](https://www.goodreports.org/static/docx/diagrams/PRISMA-2009-Flow-Diagram.docx). | 6 |
| Study characteristics | [#18](https://www.goodreports.org/reporting-checklists/prisma/info/#18) | For each study, present characteristics for which data were extracted (e.g., study size, PICOS, follow-up period) and provide the citation. | 6,7 |
| Risk of bias within studies | [#19](https://www.goodreports.org/reporting-checklists/prisma/info/#19) | Present data on risk of bias of each study and, if available, any outcome-level assessment (see Item 12). | 7 |
| Results of individual studies | [#20](https://www.goodreports.org/reporting-checklists/prisma/info/#20) | For all outcomes considered (benefits and harms), present, for each study: (a) simple summary data for each intervention group and (b) effect estimates and confidence intervals, ideally with a forest plot. | 8 |
| Synthesis of results | [#21](https://www.goodreports.org/reporting-checklists/prisma/info/#21) | Present the main results of the review. If meta-analyses are done, include for each, confidence intervals and measures of consistency. | 8 |
| Risk of bias across studies | [#22](https://www.goodreports.org/reporting-checklists/prisma/info/#22) | Present results of any assessment of risk of bias across studies (see Item 15). | 6 |
| Additional analysis | [#23](https://www.goodreports.org/reporting-checklists/prisma/info/#23) | Give results of additional analyses, if done (e.g., sensitivity or subgroup analyses, meta-regression [see Item 16]). | 8,9 |
| **Discussion** |  |  |  |
| Summary of Evidence | [#24](https://www.goodreports.org/reporting-checklists/prisma/info/#24) | Summarize the main findings, including the strength of evidence for each main outcome; consider their relevance to key groups (e.g., health care providers, users, and policy makers | 9,10,11 |
| Limitations | [#25](https://www.goodreports.org/reporting-checklists/prisma/info/#25) | Discuss limitations at study and outcome level (e.g., risk of bias), and at review level (e.g., incomplete retrieval of identified research, reporting bias). | 11 |
| Conclusions | [#26](https://www.goodreports.org/reporting-checklists/prisma/info/#26) | Provide a general interpretation of the results in the context of other evidence, and implications for future research. | 12 |
| **Funding** |  |  |  |
| Funding | [#27](https://www.goodreports.org/reporting-checklists/prisma/info/#27) | Describe sources of funding or other support (e.g., supply of data) for the systematic review; role of funders for the systematic review. | 13 |

The PRISMA checklist is distributed under the terms of the Creative Commons Attribution License CC-BY. This checklist was completed on 12. April 2021 using <https://www.goodreports.org/>, a tool made by the [EQUATOR Network](https://www.equator-network.org) in collaboration with [Penelope.ai](https://www.penelope.ai)
